# Clinical characteristics and impact of glycemic control and antifungal treatment on mortality in patients with Rhino-Orbital Mucormycosis in Mexico: A retrospective cohort study

**DOI:** 10.1101/2025.04.29.25326664

**Authors:** José Omar Jiménez-Jacinto, Bertha Beatriz Montaño-Velázquez, Andrés Tirado-Sánchez

## Abstract

**Background:** Rhino-orbital mucormycosis (ROM) is an invasive fungal infection with a high mortality rate, particularly in patients with metabolic and immunological disorders. Despite advancements in diagnosis and treatment, mortality remains significant, emphasizing the need to identify prognostic factors associated with poor outcomes.

**Objectives:** This study aimed to analyze risk factors associated with mortality in patients with ROM, with a specific focus on the impact of glycemic control, diabetic ketoacidosis (DKA), and the type of surgical approach.

**Methods:** A retrospective cohort study was conducted at a tertiary care hospital in Mexico, including 40 adult patients diagnosed with ROM between January 2019 and June 2023. Clinical, laboratory, and therapeutic variables were analyzed. Logistic regression and Kaplan-Meier survival analysis were performed to identify independent predictors of mortality.

**Results:** The overall mortality rate was 42.5%. Uncontrolled diabetes was the strongest predictor of mortality (OR 45.33, 95% CI 4.90–419.22, p <0.001). DKA was also significantly associated with increased mortality (RR 3.09, 95% CI 1.90–5.02, *p=* 0.003). Neutrophilia (≥7,500/µL) was linked to a higher risk of death (RR 6.78, 95% CI 1.77–25.87, *p* <0.001). No significant differences in survival were observed based on the type of surgical approach (*p =* 0.428).

**Conclusion:** Uncontrolled diabetes and DKA were the main predictors of mortality in patients with ROM, highlighting the critical role of metabolic management. Further studies with larger cohorts are needed to explore additional therapeutic strategies to improve survival outcomes.

**Author Summary:** Mucormycosis is a disease on the WHO priority list of fungal pathogens. It is associated with a high mortality rate, which has been reported to be as high as 80%. Diabetes mellitus is a common risk factor in mucormycosis cases. It is detected in 65%-85% of cases, especially in those with rhino-orbital mucormycosis (86.9%). It is considered a neglected disease that has increased disproportionately since the COVID-19 pandemic, although its prevalence is high, as in Iran and India. In Mexico, due to lack of knowledge among the population, it is a rare and neglected disease. We found that rhino-orbital mucormycosis, poor glycemic control and diabetic ketoacidosis were linked to higher mortality. Strict glucose control is essential. The type of surgical approach did not significantly affect the outcome. In order to refine interventions and better inform the future list of priority fungal pathogens, this study will provide robust estimates of disease burden and mortality.

## Introduction

Mucormycosis is an invasive fungal infection caused by fungi of the order *Mucorales* [1,2]. It is an opportunistic disease that predominantly affects patients with impaired immunity, such as those with uncontrolled diabetes mellitus, hematological malignancies, or those receiving immunosuppressive treatment [3,4]. The most common presentation is rhino-orbital-cerebral mucormycosis, characterized by an aggressive course and a high mortality rate [5].

Globally, differences in risk factors have been identified depending on the socioeconomic context [6]. In developed countries, mucormycosis primarily affects patients with hematological diseases and solid organ transplants [7], whereas in developing countries including Mexico, the main predisposing condition is poorly controlled diabetes mellitus, particularly diabetic ketoacidosis [8]. Despite advances in diagnostic and therapeutic strategies, mortality rate is high, with approximately 47 percent of patients dying within 3 months of the diagnosis [5], highlighting the need for studies that identify factors associated with clinical outcomes.

This study focuses on analyzing risk factors associated with mortality in patients with rhino-orbital mucormycosis (ROM), with a special emphasis on the impact of glycemic control, diabetic ketoacidosis, and the type of surgical approach. Although hyperglycemia has been described to promote the proliferation of Mucorales [9–11], and surgical intervention is considered the cornerstone of treatment [12,13], uncertainties persist regarding their influence on mortality. Identifying these factors will help clinical decision-making, optimizing treatment, and thus, lowering mucormycosis-related morbidity and mortality.

## Materials and Methods

### Study Design and Patients

A single-center retrospective cohort study was conducted in a Tertiary-care hospital in Mexico. Medical records of hospitalized patients with a confirmed diagnosis of ROM between January 1^st^, 2019 and June 30^th^, 2023 were included.

Patients were selected from medical records according to predefined inclusion criteria. Patients aged ≥18 years with confirmed diagnosis of mucormycosis via culture (growth of Mucorales on Sabouraud dextrose agar) and/or microscopy, including direct examination (broad ribbon-like aseptate hyphae) or histopathology (angioinvasion and tissue necrosis) were included.

Definitive diagnosis was made by magnetic resonance imaging in cases that could not be confirmed by microbiologic or histopathologic examination. Incomplete data on clinical records or insufficient information for variable analysis were excluded. Due to the low incidence of the disease, a consecutive case sampling method was used, including all hospitalized patients with ROM during the study period.

### Primary and secondary outcomes

The primary outcome was in-hospital mortality, defined as death following the diagnosis of mucormycosis.

Exposure variables included metabolic and immunological factors such as uncontrolled diabetes (serum glucose >180 mg/dL), diabetic ketoacidosis (DKA), neutrophilia (≥7,500 neutrophils/µL), and the type of surgical approach (external, endoscopic, or combined).

Additional variables were included to address potential confounding factors, such as systemic hypertension, bacterial or viral coinfection, the type of antifungal treatment (deoxycholate or liposomal amphotericin B), and the time from symptom onset to diagnosis.

The primary objective was to identify factors associated with mortality in mucormycosis patients. Secondary objectives included comparing survival curves based on glycemic control, evaluating the impact of surgical approach on mortality, and analyzing the relationship between neutrophilia and clinical outcomes.

### Data Collection

Data were extracted from electronic and physical medical records and recorded in a structured collection form, which included demographic information, comorbidities, laboratory parameters, diagnostic findings, administered treatments, and clinical outcomes. Data collection was conducted by trained personnel. The collected data were then encoded in an Excel 2016 spreadsheet (Microsoft Office, USA) and subsequently transferred to the analysis software for further evaluation.

As this was a retrospective study, selection bias was minimized by including all patients who met the inclusion criteria, regardless of their outcome. To reduce information bias, data were extracted exclusively from documented medical records, avoiding subjectivity derived from retrospective interviews.

### Statistical Analysis

Categorical variables were expressed as frequencies and percentages, while continuous variables were presented as mean ± standard deviation or median with interquartile range, depending on their distribution.

For bivariate analysis, categorical variables were compared using the chi-square test or Fisher’s exact test, while continuous variables were analyzed using the Student *t* test for normally distributed data and the Mann-Whitney U test for non-normally distributed data. To identify independent predictors of mortality, a binary logistic regression model was performed. Additionally, survival was assessed using the Kaplan-Meier method, comparing curves with the Log-Rank (Mantel-Cox) test. A significance level of *p=*<0.05 was considered for all statistical tests. Data analysis was performed using IBM SPSS Statistics software (IBM-Chicago, USA) for Windows version 28.0.

### Ethical Statement

The study was approved by the IMSS Research Ethics Committee (Registration Number: R-2024-3502-098) and was conducted in accordance with the ethical principles of the Declaration of Helsinki and the current Mexican regulations for health research (NOM-012-SSA3-2012). Since the study was based exclusively on clinical records without direct patient intervention, informed consent was not required. Data confidentiality was ensured through anonymization and storage in protected databases with restricted access. Missing data were also reported. This study received no external funding, and the authors declared no conflicts of interest. The methodology and reporting of results followed the STROBE guidelines for observational studies.

## Results

### Demographics and Clinical Characteristics

A total of 40 medical records of patients diagnosed with ROM were analyzed (Table 1). The mean age was 52.8 ± 13.59 years, with a slight predominance among male patients (ratio of male:female 1.1:1). One-quarter of patients were aged 60-69.

**Table 1.** Demographics and Clinical Characteristics of Patients with Mucormycosis.

| | | Mean (± SD) / Median<br>(IQR)<br>n= 40 |
| --- | --- | --- |
| Outcome<sup>†</sup> | Alive | 23 (57.5) |
|  | Deceased | 17 (42.5) |
| Age (years) | | 52.8 (± 13.59) |
| Sex<sup>†</sup> | Female | 19 (47.5) |
|  | Male | 21 (52.5) |
| Weight (Kg) | | 74.8 (± 12.89) |
| Height (m) |  | 1.61 (1.56 – 1.67) |
| BMI (Kg/m<sup>2</sup>)<sup>†</sup> | | 28.52 (± 3.15) |
|  | Normal | 6 (15) |
|  | Overweight | 21 (52.5) |
|  | Obesity grade I | 6 (15) |
|  | Obesity grade II | 1 (2.5) |
| Serum glucose (mg/dL) |  | 198.5 (133.50 – 287.50) |
| Neutrophils (mm<sup>3</sup>) |  | 7.90 (4.88 – 11.37) |
| Days from symptom onset to diagnosis (days) |  | 14.5 (9 – 21.75) |
| Comorbidities<sup>†</sup> |  | 40 (100) |
| Diabetes<sup>†</sup> |  | 35 (87.5) |
| Uncontrolled diabetes<sup>†</sup> |  | 22 (55) |
| Diabetic ketoacidosis<sup>†</sup> |  | 6 (15) |
| Systemic arterial hypertension<sup>†</sup> |  | 20 (50) |
| Neoplasia<sup>†</sup> |  | 3 (7.5) |
| Positive culture<sup>†</sup> |  | 15 (37.5) |
| Species<sup>†</sup> | *Rhizopus* | 14 (35) |
|  | *Mucor* | 1 (2.5) |
| Antifungal treatment<sup>†</sup> | Deoxycholate amphotericin B | 39 (97.5) |
|  | Liposomal amphotericin B | 1 (2.5) |
| Duration of treatment (days) |  | 19.5 (8 – 27.5) |
| Number of surgeries<sup>†</sup> | None | 1 (2.5) |
|  | 1 | 20 (50) |
|  | 2 | 14 (35) |
|  | ≥3 | 5 (12.5) |
| Type of surgical approach<sup>†</sup> | External | 19 (47.5) |
|  | Endoscopic | 10 (25) |
|  | Combined | 10 (25) |
| Days from admission to first surgery |  | 1 (1 – 2) |
| Co-infection<sup>†</sup> | Absent | 25 (62.5) |
|  | Bacterial | 14 (35) |
|  | Viral | 1 (2.5) |

<sup>†</sup>Frequency (%)

All patients had at least one comorbidity, 87.5% had pre-existing diabetes mellitus, and 62.8% of these cases had uncontrolled diabetes and 17.1% had diabetic ketoacidosis (median serum glucose level at admission was 198.5 mg/dL (range: 69–445 mg/dL). HbA1c levels were not measured in any case.

Other comorbidities include hypertension in half of the cases, and hematologic malignancy in 3 patients.

The median time from symptom onset to diagnosis was 14.5 days (range: 3–90 days). All cases were diagnosed through direct examination; however, only 37.5% had positive cultures, with *Rhizopus spp*. being the most frequently isolated species (93.3%).

Regarding treatment, 97.5% of patients received deoxycholate amphotericin B, while only one patient (2.5%) received liposomal amphotericin B. The median duration of antifungal therapy was 19.5 days (range: 3–63 days). Concerning surgical management, 97.5% of patients underwent surgery; 50% had a single surgical procedure, 35% underwent two procedures, and 12.5% had three or more. The most common surgical approach was external (47.5%), followed by endoscopic (25%) and combined (25%). The time from hospital admission to the first surgery ranged from 1 to 7 days, with a median of 1 day.

The overall mortality was 42.5% (17/40), with the highest mortality observed in patients aged 70 years or older (Figure 1). The overall survival, defined as the time from either diagnosis or treatment initiation until death from any cause, had a mean survival time of 38.5 days (95% CI: 29.8 – 47.3 days).

**Fig 1.**
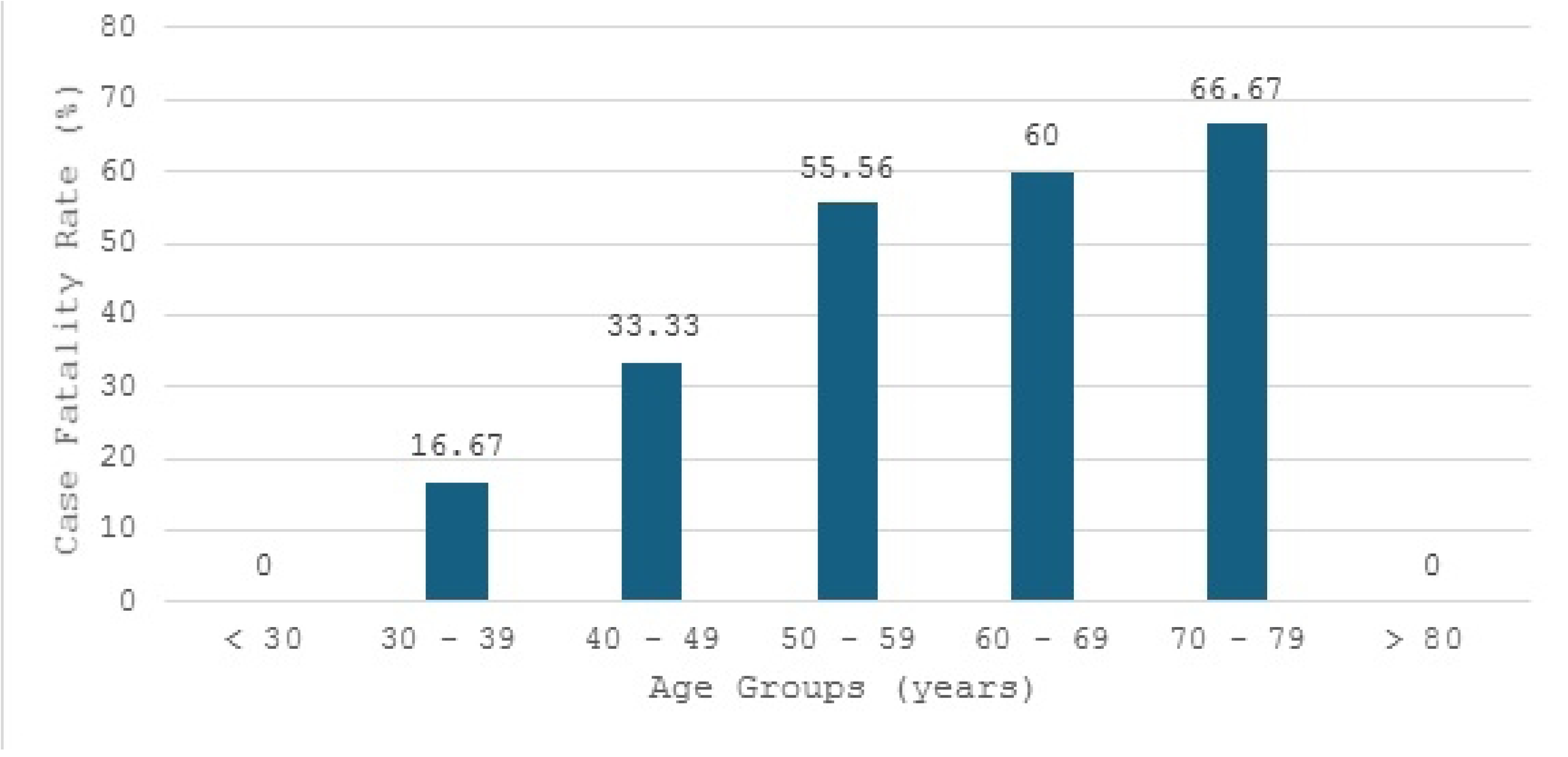
Case fatality rate by age group.

### Risk Factors Analysis

Bivariate analysis revealed that deceased patients had significantly higher serum glucose levels at admission compared to survivors (median 290 mg/dL vs. 158 mg/dL, *p=*<0.001). Similarly, neutrophil counts were significantly higher in deceased patients (median 11.45 ×10³/µL vs. 5.59 ×10³/µL, *p=*<0.001) (Table 2).

**Table 2.**
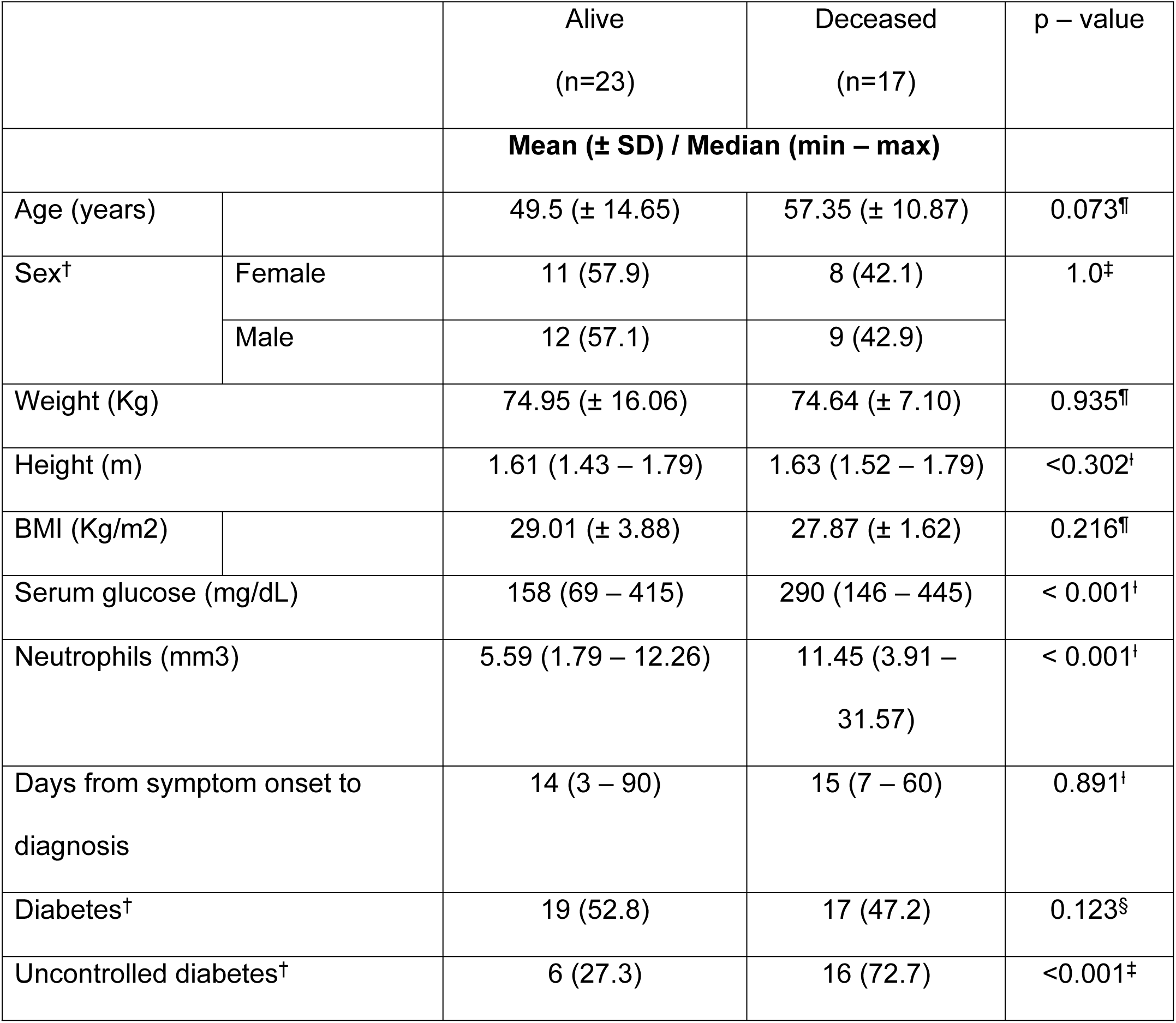
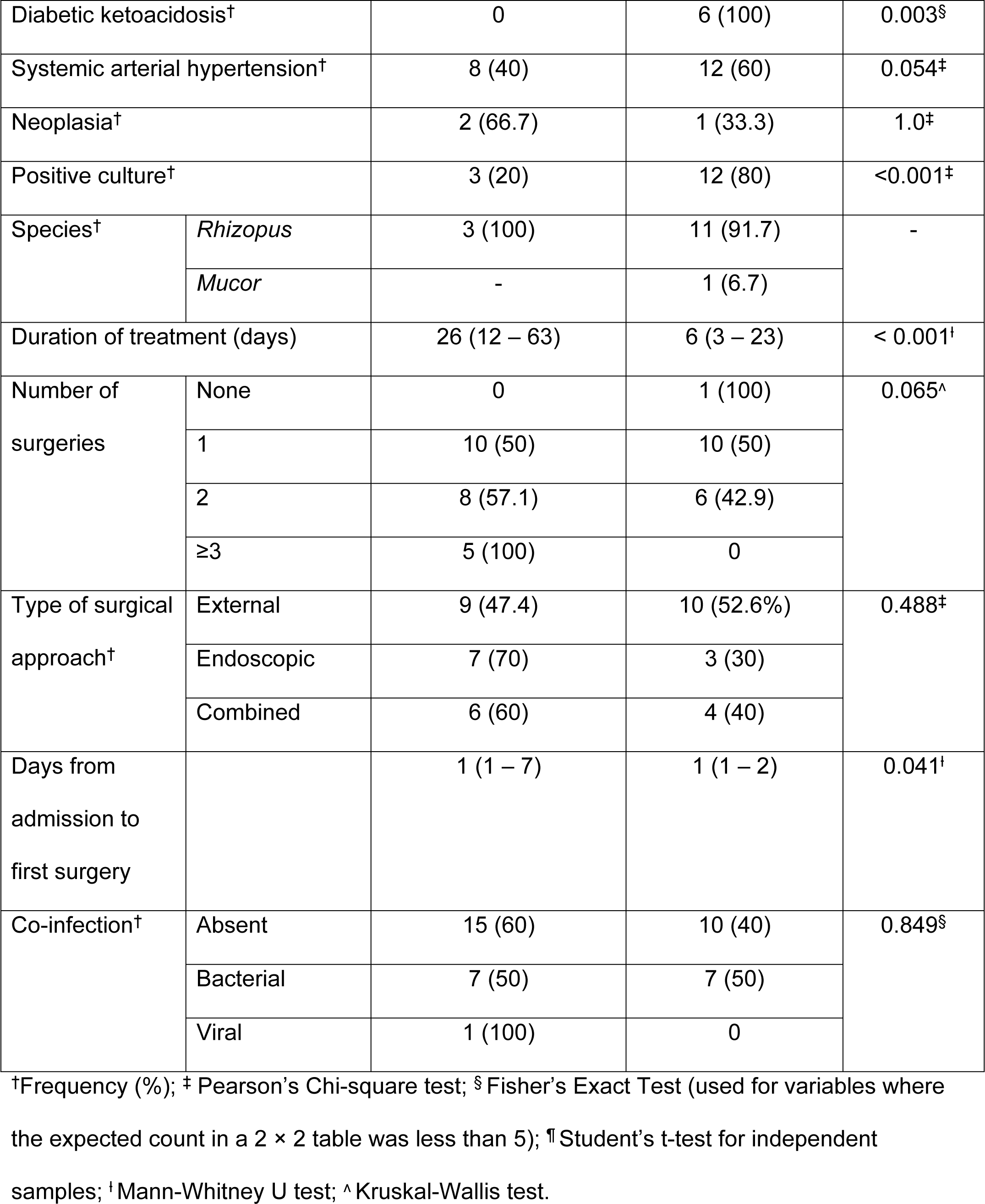
Comparison between outcomes and variables.

Uncontrolled diabetes was associated with an increased risk of mortality, with a relative risk (RR) of 13.0 (^95%^CI: 1.91–89.45, *p=*<0.001). Diabetic ketoacidosis also showed a strong association with mortality, with an RR of 3.09 (^95%^CI: 1.90–5.02, *p=* 0.003). The presence of neutrophilia (≥7,500/µL) was significantly associated with an increased risk of death (RR 6.78, 95% CI: 1.77–25.87, *p=*<0.001). No significant differences in mortality were observed according to the type of surgical approach (Table 3).

**Table 3.** Risk Factors Associated With All-Cause Mortality During a Hospital Stay.

| Variable | Relative risk (95% CI) | Phi coefficient |
| --- | --- | --- |
| Uncontrolled diabetes | 13.00 (1.91 – 89.45) | 0.676 |
| Diabetic ketoacidosis | 3.09 (1.90 – 5.02) | 0.489 |
| Age > 50 years | 2.40 (0.94 – 6.08) | 0.330 |
| Neutrophilia | 6.78 (1.77 – 25.87) | 0.615 |
| External surgical<br>approach vs. others | 1.57 (0.753 – 3.31) | 0.195 |
| Endoscopic surgical<br>approach vs. others | 0.643 (0.23 – 1.78) | 0.146 |
| Combined surgical<br>approach vs. others | 0.92 (0.39 – 2.18) | 0.029 |

In logistic regression analysis, uncontrolled diabetes was identified as the main predictor of mortality (Odds Ratio (OR) = 45.33 (95% CI: 4.90–419.22, *p=*<0.001).

Kaplan-Meier survival analysis showed that survival was worse for patients with uncontrolled diabetes compared to those with controlled serum glucose levels. The median survival for patients with uncontrolled diabetes was 8 days, whereas the median survival for the controlled glucose group was not reached, with more than 50% of patients remaining alive at the end of follow-up (*p*=< 0.001) (Figure 2). Similarly, patients with diabetic ketoacidosis had higher mortality rates than those who did not (median survival: 5.1 days versus 44.4 days (*p=*<0.001) (Figure 3).

**Fig 2.**
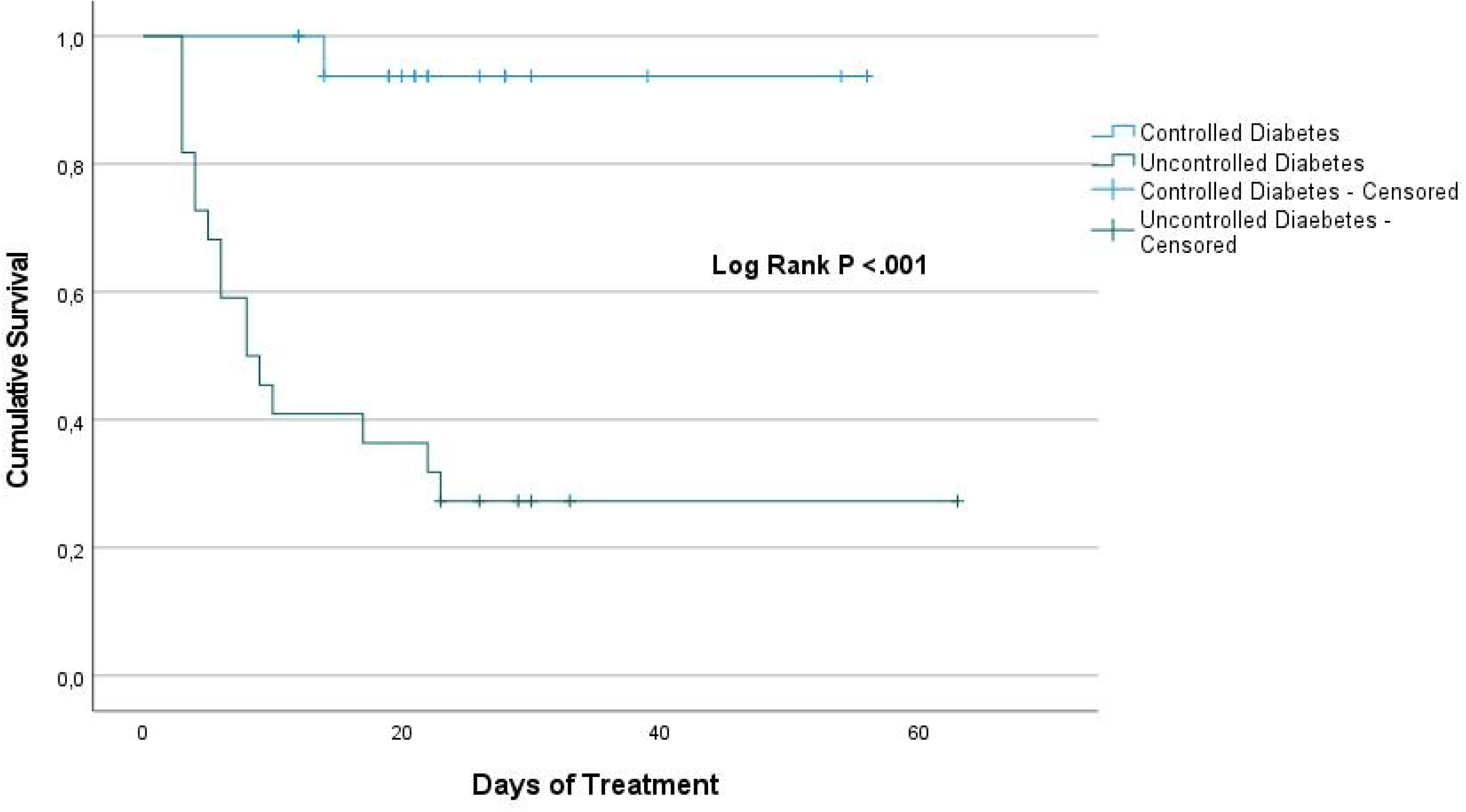
Survival probability in patients with mucormycosis with and without uncontrolled diabetes.

**Fig 3.**
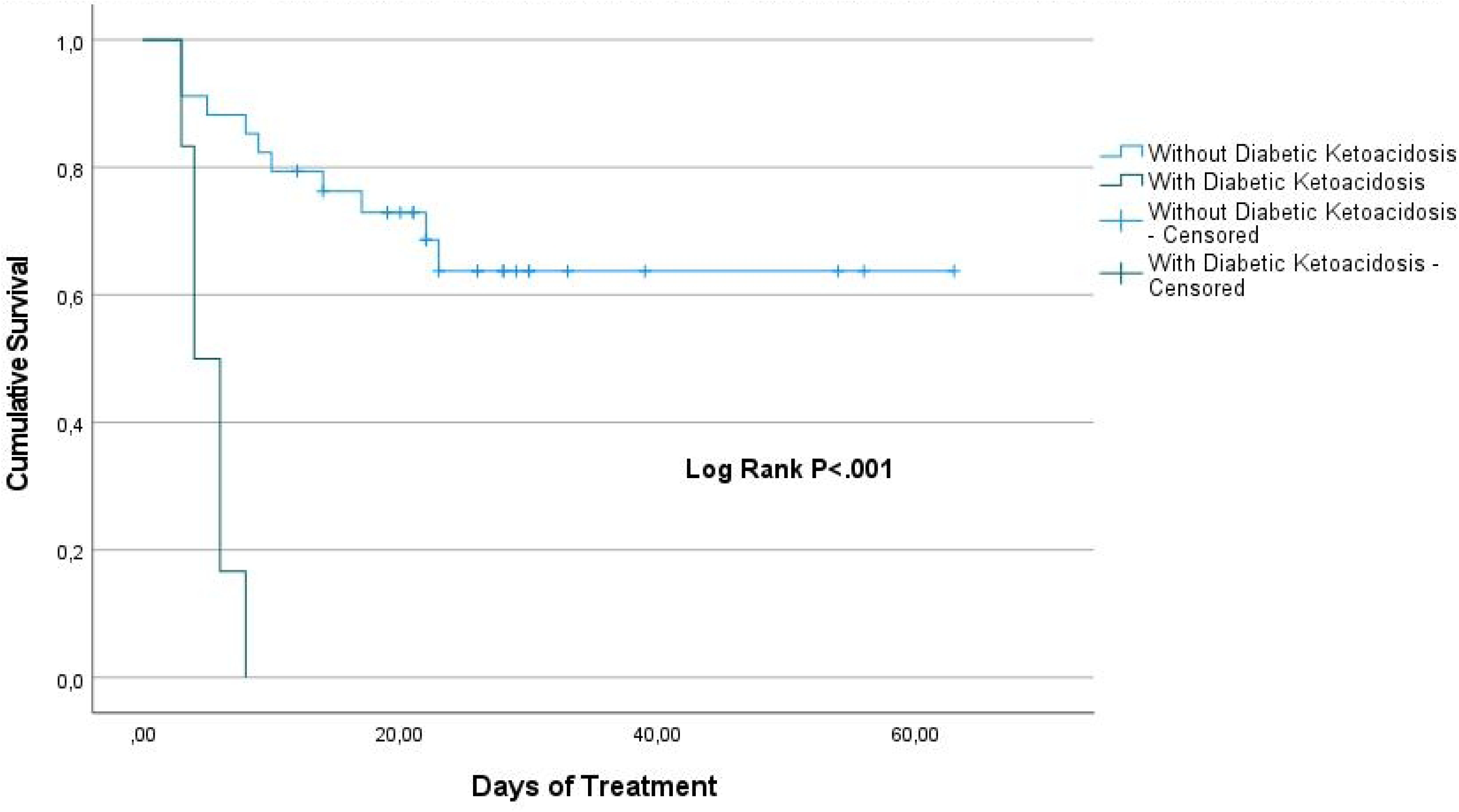
Survival probability in patients with mucormycosis with and without diabetic ketoacidosis (DKA).

Statistically significant differences in survival were not observed based on the type of surgical approach. Patients who underwent endoscopic surgery had a longer survival rate compared to those who underwent external or combined approaches. This comparison was not statistically significant, but the trend is clear (*p=*0.428) (Figure 4).

**Fig 4.**
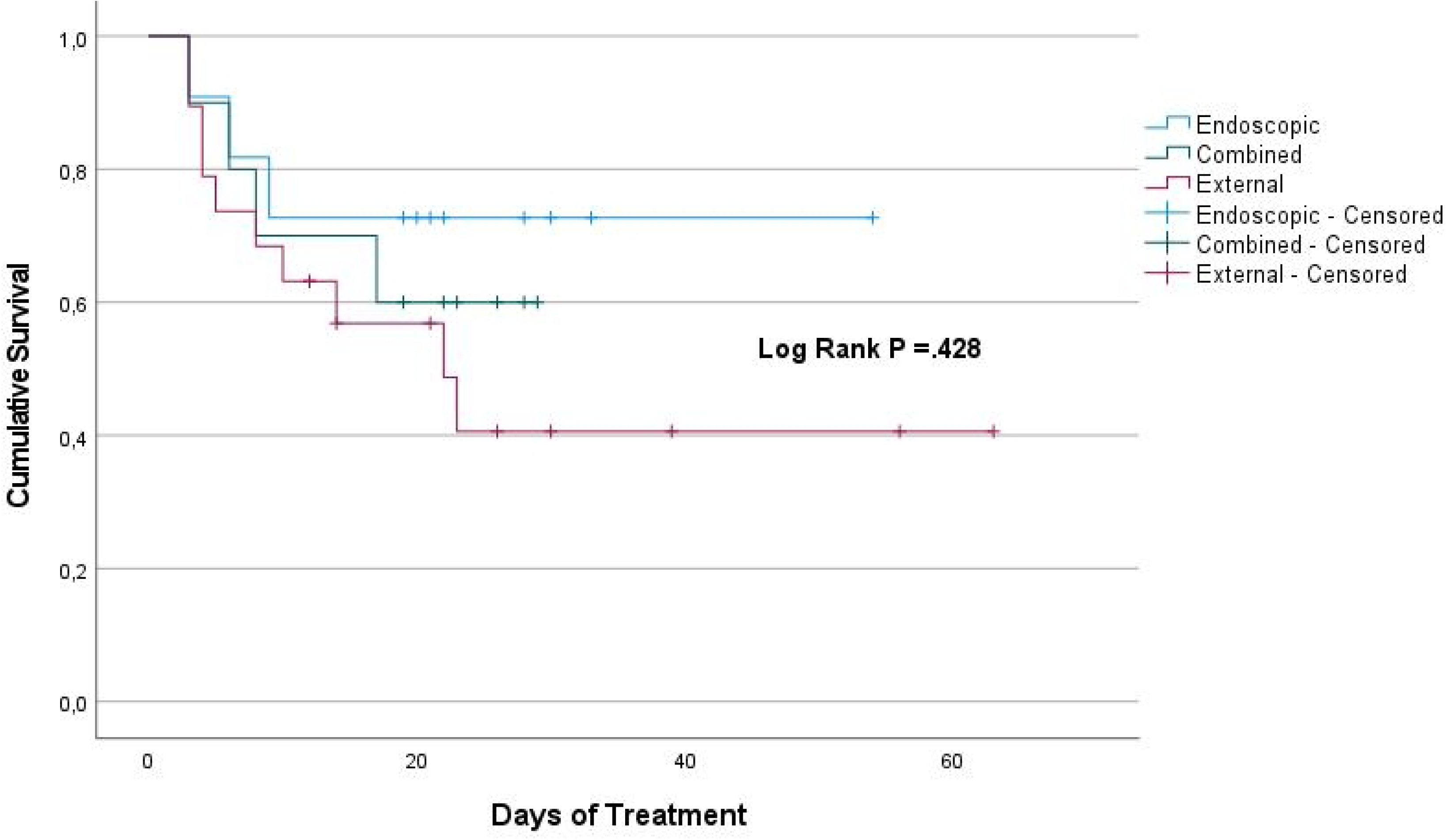
Survival probability in patients with mucormycosis by type of surgical approach.

## Discussion

ROM is an invasive fungal infection with a high mortality rate, particularly in patients with metabolic and immunological disorders [14]. This study evaluated risk factors associated with mortality in a cohort of mucormycosis patients treated at a tertiary hospital in Mexico. The results demonstrated that uncontrolled diabetes and diabetic ketoacidosis were the main predictors of mortality in this population, while the type of surgical approach was not significantly associated with clinical outcomes.

The findings of this study confirm the critical role of metabolic dysregulation in the progression of mucormycosis. Logistic regression analysis showed that hyperglycemia at hospital admission was significantly associated with increased mortality (OR 45.33, ^95%^CI 4.90–419.22, *p=*<0.001). These results are consistent with previous studies that have identified hyperglycemia as a key factor in mucormycosis progression, facilitating *Mucorales* proliferation and increasing susceptibility to angioinvasion and tissue necrosis [15–18]. Likewise, diabetic ketoacidosis had a strong impact on mortality, with a relative risk of 3.09 (^95%^CI 1.90–5.02), supporting the hypothesis that metabolic acidosis impairs the host immune response and creates a more favorable environment for fungal invasion [19,20].

Moreover, neutrophilia was also associated with an increased risk of mortality, potentially reflecting an exacerbated inflammatory response in patients with poorer prognosis. While neutropenia is a well-established risk factor for mucormycosis [21,22], previous studies suggest that in hyperglycemic patients, neutrophil dysfunction may lead to an ineffective immune response against the fungus, thereby increasing mortality [16].

In hyperglycemic and ketoacidotic conditions—common in diabetic patients—neutrophil function is significantly impaired. Their migration toward Mucorales hyphae and ability to eliminate them are markedly reduced, which may explain why patients with elevated neutrophil counts still fail to control the infection, ultimately leading to worse outcomes [16,23].

Our findings suggest that, beyond hyperglycemia, neutrophilia may reflect an excessive yet ineffective inflammatory response that does not necessarily improve patient prognosis. Instead, high neutrophil counts may be linked to increased production of reactive oxygen species (ROS) and proinflammatory cytokines, which can exacerbate tissue damage and disease progression rather than effectively containing the infection. This is consistent with prior research indicating that diabetes-associated neutrophil dysfunction results in chronic inflammatory activation, excessive ROS production, and impaired pathogen clearance, further contributing to increased susceptibility and mortality in severe infections [24,25].

Regarding surgical treatment, no significant differences in mortality were observed among different surgical approaches (*p=*0.428). While previous studies have suggested that endoscopic surgery may be associated with better outcomes due to lower morbidity and faster recovery [26–29], no significant impact was observed in this cohort. However, this could be attributed to the sample size or the selection of more advanced cases for more invasive surgical approaches.

### Clinical Implications

The findings of this study reinforce the importance of metabolic control in patients with ROM. Given that hyperglycemia and diabetic ketoacidosis had a significant impact on mortality, optimizing glycemic control should be a priority in these patients. This includes early monitoring and aggressive management of dysglycemia from hospital admission. Additionally, these results suggest that surgical strategies should be individualized, with other factors, such as fungal burden and the patient’s immune status, potentially influencing outcomes beyond the type of surgical approach.

### Future Directions

Given the significant impact of glycemic dysregulation on mucormycosis progression, future studies should focus on interventions aimed at improving early metabolic control in these patients. Clinical trials evaluating intensive glucose management strategies and acid-base balance correction could provide stronger evidence on how to mitigate the impact of these factors on mortality. Furthermore, multicenter studies with larger sample sizes would allow for a more precise assessment of the role of different surgical techniques in survival.

### Strengths and Limitations

One of the strengths of this study is its design based on real-world clinical data from hospitalized patients at a reference center, allowing for a detailed evaluation of risk factors associated with mortality. Additionally, the inclusion of a retrospective cohort with an adjusted logistic regression analysis strengthens the validity of the findings.

However, the study presents some limitations. The sample size was relatively small, which may have limited the ability to detect significant associations in some variables, particularly the impact of the type of surgical approach. Furthermore, as a retrospective study, the quality of the data depended on documentation in medical records, potentially introducing information bias. Finally, as a single-center study, the results may not be fully generalizable to other populations with different access to medical resources and therapeutic strategies.

## Conclusions

This study identified uncontrolled diabetes and diabetic ketoacidosis as the main factors associated with mortality in patients with ROM, whereas the type of surgical approach did not significantly influence outcomes. Hyperglycemia was associated with a substantial increase in mortality, highlighting the importance of strict metabolic control as an integral component of mucormycosis management.

These findings underscore the need for optimized glycemic control in mucormycosis patients and for continued exploration of therapeutic strategies that may improve survival in this high-risk population. Additional studies with larger sample sizes and a prospective design are required to validate these results and evaluate novel therapeutic strategies aimed at reducing the high mortality associated with this invasive fungal infection.

## Data Availability

All data produced in the present study are available upon reasonable request to the corresponding author.

